# Aortic root anatomy and impact on new-onset conduction abnormalities after transcatheter aortic valve implantation

**DOI:** 10.1101/2024.08.30.24312869

**Authors:** Habib Layoun, Joseph Kassab, Michel Chedid El Helou, Joseph El Dahdah, Odette Iskandar, Maryam Muhammad Ali Majeed Saidan, Abdelrahman Abushouk, Toshiaki Isogai, Grant Reed, Rishi Puri, Oussama M. Wazni, Amar Krishnaswamy, Serge Harb, Samir Kapadia

**Author notes:** Address for correspondence: Samir R. Kapadia, MD, Chairman, Department of Cardiovascular Medicine Cleveland Clinic Foundation, 9500 Euclid Avenue, Cleveland, OH 44195. Disclosures: None.

## Abstract

**Background:** Angulation of virtual basal ring (VBR), also known as aortic annulus, in relation to sino-tubular joint (STJ) may lead to greater exposure of implanted stent to the conduction system, consequently increasing the risk of LBBB.

**Objective:** To measure VBR-STJ angle and explore its impact on the development of LBBB post TAVR.

**Methods:** Patients undergoing TAVR using the Sapiens 3 valve between 2016 and 2021, without pre-TAVR conduction anomalies were included. The angle between the VBR and the ascending aorta was measured as the angle between the VBR plane and the plane of the STJ on cardiac CT, along with the annulus dimensions. TAVR implantation depth was measured on intra-procedural fluoroscopy images.

**Results:** 1204 patients were included, with 145 having new-onset LBBB. The VBR-STJ angle was significantly greater in the new-onset LBBB group (7.3 ± 4.7 vs 5.9 ± 4.6, p=0.002), and the difference in implantation depth between the levels of right and none coronary cusp (RCC and NCC) was significantly correlated with the VBR-STJ angle (r=0.1, p=0.03). This angle was further associated with new-onset LBBB after adjustment to patient and procedural characteristics (OR 1.08 CI: [1.04, 1.13], p<0.001).

**Conclusion:** Patients developing LBBB have larger VBR-STJ angle which was associated with greater depth of implantation of the TAVR valve below the RCC compared to the NCC. Precise understanding of the aortic root anatomy can help to predict onset of LBBB which in turn can inform decision making regarding optimal way of treating aortic stenosis and may improve procedure planning.

**Condensed abstract:** This study explores the impact of the virtual basal ring (VBR) to the sino-tubular junction (STJ) angle on the development of new-onset left bundle branch block (LBBB) post TAVR. The angle was measured in 1204 patients who underwent TAVR, with 145 developing LBBB post TAVR. The measured angle was significantly higher in patient with new-onset LBBB and was independently correlated with new-onset LBBB. These findings emphasize the importance of understanding the aortic root anatomy to improve TAVR planning and reduce the incidence of LBBB.

## Introduction

Recent advancements in transcatheter aortic valve replacement (TAVR) have led to a significant increase in the number of aortic valve replacements (1) due to its safety and effectiveness. However, new-onset left bundle branch block (LBBB) remains one of the most frequent conduction disturbances following TAVR (2), with resulting adverse outcomes (3).

Due to the proximity of the superior fascicle of the left bundle branch to the aortic root, the conduction system remains at a high risk of disruption during TAVR, especially while it passes in the membranous septum of the left ventricular outflow tract (LVOT), right below the right coronary artery leaflet (4). Several procedural factors have been linked to a higher risk of LBBB development, such as implantation depth of the TAVR valve, overstretching of the LVOT with larger prosthesis-to-LVOT ratio and the use of self-expanding TAVR valves (2).

Since the initial description by Piazza et al of the virtual basal ring (VBR) and sino-tubular joint (STJ) planes (5), its relationship to outcomes have not been explored in detail. Although they are depicted as parallel planes, the characterization of the angle between the two planes has not been studied in the TAVR population. Further, scarce literature exists on the importance of this anatomical feature in relation to conduction system abnormalities in patients undergoing TAVR. Therefore, we sought to measure the angle between the STJ and the VBR on pre-TAVR cardiac computed tomography (CCT), and investigate the potential impact of higher angles on the development of new-onset LBBB.

## Methods

We conducted a retrospective case-control study, including patients ≥18 years old, who underwent TAVR with a Sapien 3 (S3) valve (Edwards Lifesciences, Irvine, California) at the Cleveland Clinic between January 2016 and December 2019. Patients with pre-existing cardiac pacing devices or wide QRS (≥120 ms) on pre-TAVR electrocardiogram and those who had implanted valves other than the S3 were excluded. Patients included in the final analysis were then divided into those who developed new onset LBBB after the procedure and those who did not. The Cleveland Clinic Institutional Review Board approved this study.

### Electrocardiogram (ECG)

For all study participants, 12-lead ECGs were taken at a speed of 25 mm/s and calibration of 1 mV/mm, at two time points: pre-TAVR and on the first day post-TAVR. Two experienced cardiologists reviewed and interpreted all ECG tracings, following the standard definitions and guidelines recommended by the American Heart Association, American College of Cardiology Foundation, and Heart Rhythm Society (6,7).

### Baseline characteristics

Patients’ characteristics including demographics, cardiovascular comorbidities and risk factors were extracted from the electronic medical records.

### Cardiac CT (CCT)

The following measurements were manually performed by the authors on pre-TAVR CCT. The virtual basal ring (VBR), also known as the aortic annulus, was first identified on pre-TAVR CCT on short-axis view by identifying the nadir for each aortic valve leaflet attachment. A virtual centerline was then drawn in space, perpendicular to the VBR, as seen on both coronal and sagittal views (Figure 1). The STJ was then identified and similarly, a centerline was drawn perpendicular to its plane. The angle between both planes was then measured as the angle in space between the two virtual centerlines. VBR cross sectional area was also measured and the implanted prosthesis valve diameter-to-VBR area ratio was calculated.

**Figure 1:**
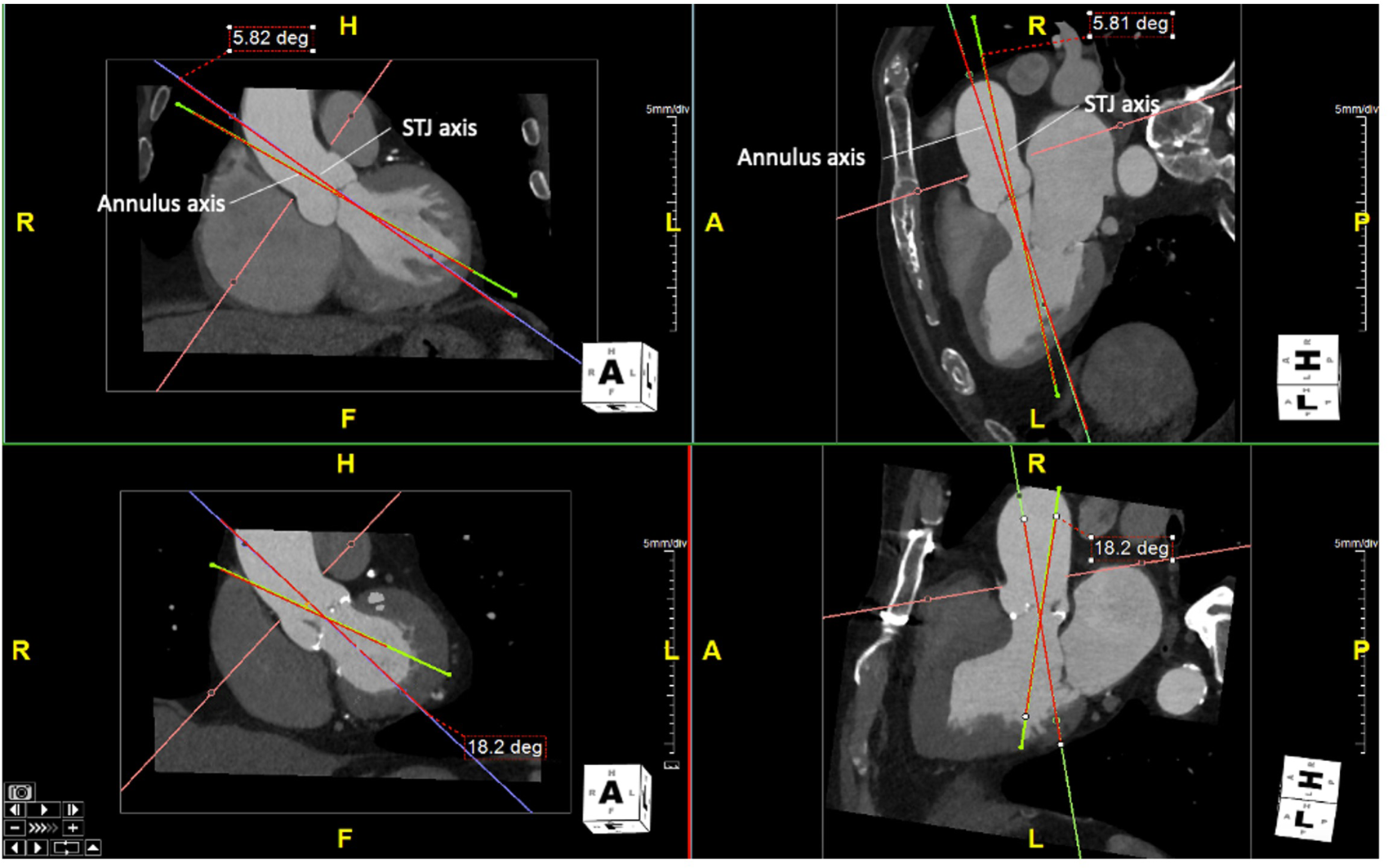
Measurement of the VBR angle defined as the angle between the axis of the STJ plane and the VBR plane, in a patient without LBBB (upper panels) and a patient who developed post-TAVR LBBB (lower panels)

We further measured aortic valve calcium score using the Agatston score. To adjust for contrast enhanced studies, a region of interest was placed in the ascending aorta and the mean attenuation value along with the standard deviation were recorded. We then calculated the threshold for calcium detection as mean attenuation + 2SD (8).

### Fluoroscopy

Intra-procedural fluoroscopy recordings were used to measure valvular implantation depth on right anterior oblique (RAO) view. The RAO view was selected for its ability to provide optimal visualization of the right coronary cusp (9). The implantation depth was measured as the distance between the bottom of the left (non-coronary sinus) and right (right coronary sinus) borders of the aortic valve sinus, and the stent frame (Figure 2). To further assess the association of VBR-STJ angulation with prosthetic valve implantation, we measured the difference in implantation depth between the right coronary sinus side and the non-coronary sinus side, and tested the correlation of this difference with the VBR-STJ angle.

**Figure 2:**
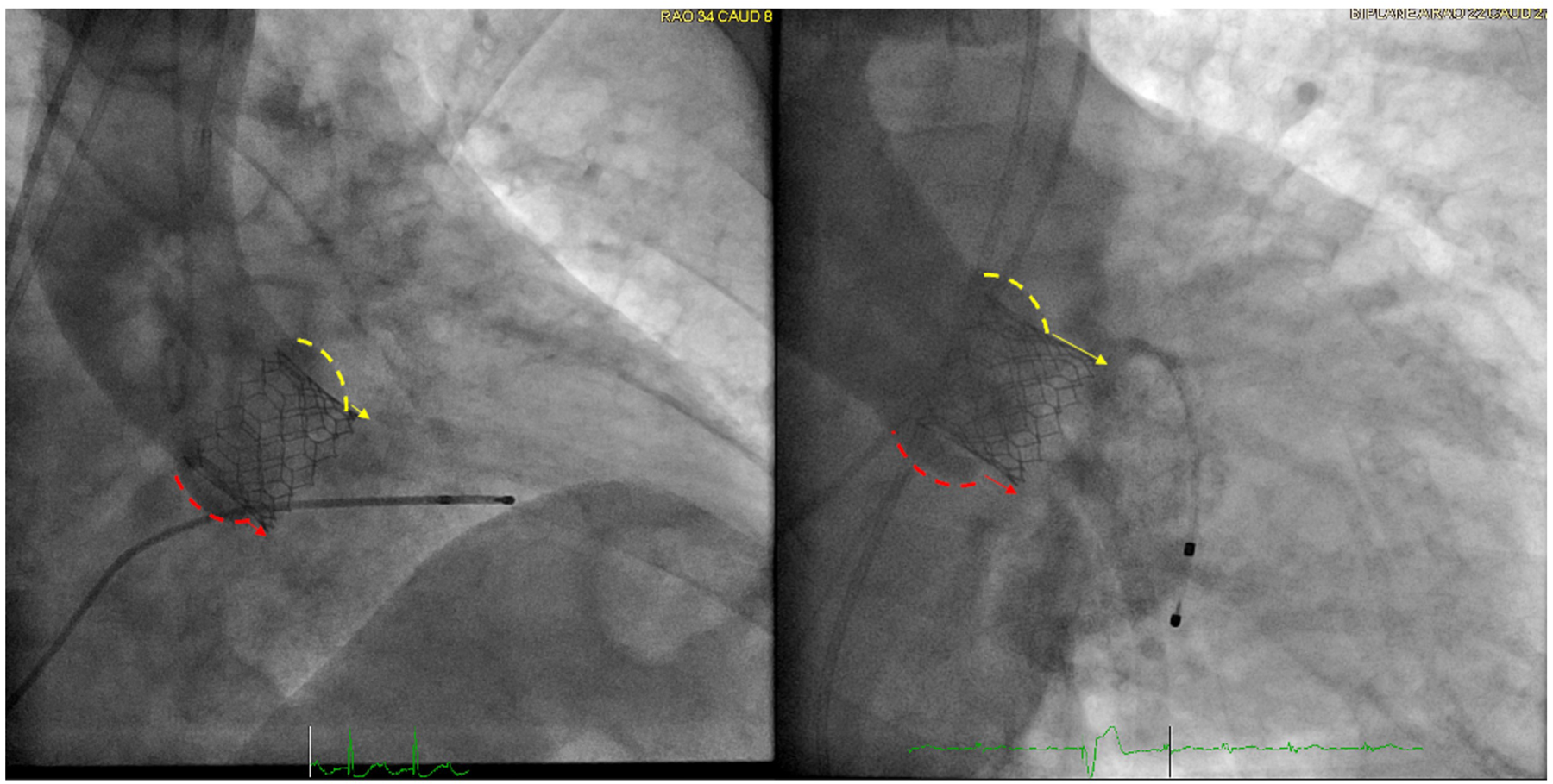
Right anterior oblique fluoroscopic images in a patient who did not develop LBBB (left panel) vs a patient who developed LBBB post-TAVR (right panel). Implantation depth of the TAVR valve was measured as the distance between the lower border of the right coronary cusp (yellow) and the non-coronary cusp (red)

### Statistical analysis

Baseline characteristics were compared between study groups using a 2-sided Student t test or Mann-Whitney U test for continuous variables and analysis of variance for categorical variables. Continuous variables are represented as mean (standard deviation) or median (interquartile percentage), and categorical variables are reported as proportions.

Association of procedural and structural variables with the onset of LBBB post TAVR was assessed using a multivariable binary logistic regression, with the presence or absence of LBBB as the dependent variable. Variables included in the model were the VBR-STJ angle, the valve size-to-VBR area ratio and the implantation depth. Pearson’s correlation test was used to compute the correlation between the VBR-STJ angle and the differences in implantation depth at the levels of right coronary cusp minus non-coronary cusp. All statistical analyses were completed using R software (R Foundation for Statistical Computing, Vienna, Austria).

## Results

### Patient population

Table 1 summarizes the clinical characteristics of the included patient population. 1204 patients were included, and 145 patients had new-onset LBBB. Mean age was and 79 years old and 46% of the population were females. Patients who developed new-onset LBBB had similar age, gender and cardiovascular comorbidities to those who did not develop LBBB. However, they had significantly higher prevalence of prior coronary artery bypass surgeries (36.5% vs 24.1%, p<0.001). Out of the 1204 included patients, 8 patients ended-up requiring permanent pacemaker implantation later on follow-up, 3 of which had developed new-onset LBBB post-TAVR.

**Table 1.** Clinical characteristics of included patients.

| Variable | No LBBB<br>(n=1059) | New-onset LBBB<br>(n=145) | p-value |
| --- | --- | --- | --- |
| Age, years | 79.1 ± 9.3 | 79.6 ± 9.5 | 0.78 |
| Female gender (%) | 487 (46.0) | 66 (45.5) | 0.98 |
| PCI (%) | 266 (25.1) | 38 (26.2) | 0.86 |
| CABG (%) | 255 (24.1) | 48 (33.1) | 0.03 |
| Myocardial infarction (%) | 213 (20.2) | 34 (23.4) | 0.42 |
| Stroke (%) | 134 (12.7) | 24 (16.6) | 0.24 |
| Hypertension (%) | 950 (89.7) | 128 (88.3) | 0.68 |
| Diabetes (%) | 366 (34.6) | 54 (37.2) | 0.59 |
| Dialysis (%) | 31 (2.9) | 3 (2.1) | 0.75 |
| Atrial fibrillation (%) | 399 (37.7) | 57 (39.3) | 0.78 |

### Virtual basal ring anatomy

The mean VBR diameter of the population was 5.4 ± 2.0 cm^2^ with a mean prosthesis to VBR ratio of 5.1 ± 1.3 mm/cm^2^. Patients who developed LBBB had similar VBR area and prosthesis-to-VBR ratio compared to those who did not develop LBBB post-TAVR (5.1 ± 1.3 vs 5.2 ± 1.3 mm/cm^2^, p=0.36). However, the VBR-STJ angle averaged at 6.0 ± 4.6 degrees, and was significantly higher in patients who had new-onset LBBB (7.3± 4.7 vs 5.86 ± 4.6 degrees, p=0.002) (Table 2).

**Table 2.** Cardiac CT- and fluoroscopy-derived measurements.

| Variable | No LBBB<br>(n=1059) | New-onset LBBB<br>(n=145) | p-value |
| --- | --- | --- | --- |
| **Cardiac CT** |  |  |  |
| VBR-STJ angle (degrees) | 5.86 (4.6) | 7.3 (4.7) | **0.002** |
| Annular area (cm<sup>2</sup>) | 5.40 (2.0) | 5.2 (2.0) | 0.60 |
| Prosthesis diameter to annular area ratio<br>(mm/cm<sup>2</sup>) | 5.12 (1.3) | 5.24 (1.3) | 0.36 |
| Aortic valve calcium score | 955.5 ± 818 | 1057.3 ± 806 | 0.22 |
| **Fluoroscopy** |  |  |  |
| Depth at non-coronary cusp | 0.24 (0.2) | 0.27 (0.2) | 0.07 |
| Depth at right-coronary cusp | 0.39 (0.2) | 0.44 (0.2) | **0.02** |

### Prosthesis implantation depth

The average implantation depth, as measured on fluoroscopy, was 0.4 ± 0.2 cm at the level of the RCC and 0.24 ± 0.19 at the level of the NCC. Patients who developed LBBB had significantly higher implantation depth at the level of the RCC compared to those who did not develop LBBB (0.44 ± 0.2 vs 0.39 ± 0.2, p=0.02), however, a similar depth was observed at the level of the NCC (0.27 ± 0.2 vs 0.24 ± 0.2, p=0.07). The difference in implantation depth between the levels of RCC and NCC was significantly correlated with the VBR-STJ angle (r=0.1, p=0.03).

### Association of VBR-STJ angle with left bundle branch block

The CT-derived VBR-STJ angle was independently associated parameter with the onset of LBBB on multivariable logistic regression (OR 1.08, p<0.001), adjusted to the prosthesis diameter-to-VBR area ratio, implantation depth and aortic valve calcium score (Table 3).

**Table 3.** Multivariable logistic regression for the association of procedural variables with the onset of CA post-TAVR.

| **Variable** | **OR (95% CI)** | **p-value** |
| --- | --- | --- |
| **VBR-STJ angle, degrees** | 1.08 (1.04, 1.13) | **<0.001** |
| **Prosthesis diameter to annular area ratio** | 1.12 (0.94, 1.33) | 0.13 |
| **Depth at non-coronary cusp side, mm** | 2.64 (0.75, 9.22) | 0.22 |
| **Depth at right-coronary cusp side, mm** | 2.16 (0.66, 6.89) | 0.20 |
| **Aortic valve calcium score, AU** | 0.99 (0.98, 1.01) | 0.22 |

## Discussion

Our study is the first to evaluate and report the association of angulation within the aortic root structures, namely the VBR and STJ, and its impact on valvular implantation during TAVR procedure. This angulation results in greater implantation depth from the right coronary cusp side, which in turn leads to exposure of the conduction system to the prosthetic valve with subsequent new onset LBBB (central figure).

Although various anatomical, procedural, and functional factors have been linked to the emergence of new onset LBBB following TAVR (2,10,11), it persists as the predominant procedural complication, underscoring an existing gap in addressing this complication. Furthermore, several studies have reported significant association with as sudden cardiac death, ventricular remodeling, and heart failure hospitalization (12–15).

During the procedure, the prosthetic valve is advanced through the ascending aorta towards the VBR, and its implantation typically follows the trajectory of the ascending aorta, parallel to the STJ plane. The VBR and STJ planes are classically depicted as parallel structures, yet, whenever an increased VBR-STJ angle exists, the angle between the bottom border of the stent and the VBR would also increase, resulting in a skewed implantation at the level of the VBR.

Several studies have examined the relationship between implantation depth and the development of LBBB after TAVR, both with balloon-expanding (BE) and self-expand (SE) valves (16,17). In our study, we observed a higher incidence of LBBB in patients with greater implantation depth at the level of the right coronary cusp which was weakly yet significantly correlated with the VBR-STJ angle due to the skewed implantation, which was best visualized on the RAO view (9). This finding aligns with recent studies delineating the course of the superior fascicle of the left bundle, which traverses the nadir of the right coronary cusp, rendering it particularly susceptible to rhythm disturbances (4). Additionally, other factors have been linked to the onset of LBBB post-TAVR, such as pre-existing conduction anomalies (18), valve size (19), self-expanding valves (10), overexpansion of the native aortic annulus (2) and higher aortic valve calcium score (10). To ensure clarity in our analysis, patients with pre-existing conduction disorders and those receiving self-expanding valves were deliberately excluded from our study cohort. The remaining variables, including the VBR-STJ angle, were subjected to logistic regression analysis. Our findings demonstrated that the VBR-STJ angle emerged as an independent predictor associated with the development of LBBB following TAVR. Further investigation is warranted to corroborate these findings and ascertain whether procedural adjustments based on the VBR-STJ angle could mitigate the incidence of LBBB post-TAVR, ultimately enhancing patient outcomes.

## Limitations

It is important to acknowledge several limitations of this analysis. On one hand, it is a retrospective study conducted at a single high-volume center, and only includes S3 recipients, which may restrict the generalizability of our findings. On a second hand, the use of automated measurements of PR interval and QRS duration by a computer-based algorithm may have introduced some slight variability into the results.

## Conclusion

In conclusion, our study found that patients with new-onset LBBB had higher VBR-STJ angles which was associated with deeper implantation of the prosthetic valve below the RCC. These findings suggest that a greater understanding of the aortic root anatomy could aid in procedure planning and patient selection for TAVR procedure to potentially reduce the incidence of LBBB.

## Data Availability

Data referred to in the manuscript is not available for sharing per the institutional policies

**Central Figure:** Patients who experienced new onset post-TAVR LBBB had increased aortic annular angulation, causing a greater exposure of the conduction system to the TAVR valve at the level of the right coronary cusp. STJ: Sino-tubular joint; NCS: Non-coronary sinus; RCC: Right coronary sinus; VBR: Virtual basal ring.

